# Associations of menstrual health with school absenteeism and educational performance among Ugandan secondary school students: A longitudinal study

**DOI:** 10.1101/2025.06.03.25328933

**Authors:** Christopher Baleke, Levicatus Mugenyi, Kate A Nelson, Katherine A Thomas, Denis Ndekezi, Jonathan Reuben Enomut, Connie Alezuyo, John Jerrim, Helen A Weiss

**Author notes:** Correspondence to be addressed to: Christopher Baleke.

## Abstract

**Background:** Relatively few studies have quantified the amount of school missed due to poor menstrual health, or the impact of menstrual health on educational performance.

**Methods:** We analysed baseline and longitudinal data from participants in a cluster-randomised trial of a menstrual health intervention in 60 Ugandan secondary schools. We measured school absenteeism as the self-reported number of days absent due to menstruation per month and assessed educational performance using examination results, at baseline and endline. We estimated adjusted incidence rate ratios (aIRR) for associations with school absenteeism, using negative binomial regression adjusted for school-level clustering. We estimated adjusted standardised mean differences (aSMD) in examination scores using mixed-effects linear regression.

**Results:** At baseline, 3312 participants reported menstruating in the past 6 months (mean age 15.6 years). Of these, 323 (9.8%) participants reported missing at least one day of school per month due to menstruation and 1286 (38.8%) missed at least one day per month for any reason. Of the 1192 participants in the control arm seen at endline, 135 (11.3%) missed at least one day due to menstruation (mean days missed=0.30 per month (95%CI 0.27-0.34)). Menstrual-related absenteeism and poorer examination performance at endline were both associated with multiple dimensions of menstrual health (use of inadequate menstrual materials, negative menstrual attitudes, unmet menstrual practice needs, and experience of menstrual teasing). In addition, absenteeism (due to menstruation and overall) was associated with menstrual pain.

**Conclusion:** Among Ugandan students, multiple dimensions of menstrual health are associated with school absenteeism and educational performance.

## 1. Introduction

Menstrual health (MH) is defined as complete physical, mental, and social wellbeing in relation to the menstrual cycle (1) . Poor menstrual health is prevalent in low- and middle-income countries (LMICs), and a review of qualitative studies in LMICs found that multiple aspects of menstrual experience contribute to school absenteeism, including inadequate menstrual materials, poor menstrual knowledge and confidence, menstrual pain, and inadequate infrastructure for managing menstruation in schools (2).

A widely-cited statistic that “one in 10 school-age girls in Africa misses school or drops out for reasons related to her period” is not evidence-based (3, 4). A systematic review of 15 quantitative studies on school absenteeism during menstruation in sub-Saharan Africa found that 31% (95%CI 24-39%) of female students reported school absenteeism during menstruation either at last menstrual period (LMP) or over a longer duration, and that menstruation also affected concentration, academic performance, and participation in sports (5). However, the number of school days missed were not quantified and there was a high degree of unexplained heterogeneity between studies (I^2^=97.8%) with prevalence of absenteeism due to menstrual pain ranging from 6% in Ghana to 65% in Ethiopia. The reference period for measurement of school absenteeism varied, with some using LMP and others using longer durations - this may partly be responsible to variations in prevalence. A recent multi-country study of data from 673,380 women and girls aged 15-49 in 47 countries found that the proportion reporting menstrual-related school absenteeism during LMP was 15.0% (95%CI 12.7-17.3%), with the highest absenteeism among 15-19 year olds (17.7%, 95%CI 15.1-20.3%) (6). There is mixed evidence of an impact of menstrual health interventions on school absenteeism. A recent systematic review identified nine relevant studies in LMICs, of which six found evidence of a positive effect of an intervention on absenteeism, but the studies deemed to be of highest quality tended to find no effect (7). Reasons for the lack of knowledge on the extent to which menstruation affects school attendance include a lack of appropriate and cross-validated absenteeism measures (8), a lack of detail on whether absence was due to menstruation or non-menstrual reasons (such as general illness or lack of ability to pay school fees), and the time period over which absence was assessed (9).

A systematic review and meta-analysis of the association of menstrual pain (dysmenorrhea) with absenteeism among adolescents and young women identified 38 studies (including 23 from LMIC), and found that 20.1% of participants reported absence from school or university due to dysmenorrhea (95% CI 14.9–26.7) (10). A systematic review of menstrual experiences among university students identified 74 quantitative studies globally (26 in Africa), with reported consequences of menstruation including academic performance, mainly due to menstrual pain or heavy menstrual bleeding (11). There is relatively little, and inconclusive evidence on the impact of menstruation on broader educational issues such as self-esteem, participation in school and educational performance (12, 13). In the review among university students, few studies provided details of how the educational outcomes were measured, and none used extant data (official examination results and attendance records) to validate self-reported measures (11).

Formative research in Ugandan secondary school students has shown strong evidence that participants missed school more frequently during menstruation than on other days, based on prospective daily diaries (28.4% of schooldays were missed during menstruation compared with 6.5% when not menstruating; p<0.001) (14). Our recent cluster-randomised trial evaluating the impact of a multi-component menstrual health intervention showed strong evidence of an intervention effect on multiple dimensions of menstrual health over a one-year period (15), but little evidence of an impact on school absence (measured using prospectively-collected daily diaries) during menstruation (11.2% vs 13.5% days missed; adjusted odds ratio (aOR)=0.81, 95%CI 0.62, 1.05). There was no evidence of an impact of the intervention on examination performance (adjusted mean difference (aMD)=0.06, 95%CI -0.12, 0.24) or on school absence overall (10.5% days missed in both arms).

The aim of this paper is to quantify factors associated with menstrual-related school absenteeism among Ugandan secondary school students, using data from the MENISCUS trial. These findings will add to the knowledge based on the impact of multiple dimensions of menstruation on the number of school days missed due to menstruation, and on examination performance.

## 2. Methods

### 2.1 Study Setting

The study was conducted in Kalungu and Wakiso districts of Uganda. Wakiso district is the largest and largely urban district in central Uganda with an estimated population of 3.4 million people (16) . In Wakiso, 17% of residents were aged 10-17 years, of whom 49% were attending secondary schools (17). Kalungu is a rural district with population size 183,232 (2014 census). In Kalungu, 23% were aged 10-17 years, of whom 36% were enrolled in secondary school (18).

### 2.2 Data source

This study uses baseline and endline survey data from the MENISCUS trial (“Menstrual health interventions, schooling, and mental health problems among Ugandan students”) which aimed to evaluate whether a multi-component menstrual health intervention improves education, health and wellbeing among female secondary school students in Central Uganda. Details of the trial design and results have been published previously (15, 19). In brief, the trial was a parallel-arm, cluster-randomised controlled trial in 60 schools with schools randomised 1:1 to either immediate or delayed intervention delivery.

### 2.3 Sampling of schools

All government schools and a random selection of private schools were contacted through phone calls and assessed for eligibility using the government’s 2019 master list of Education Institutions. Schools were eligible if they were mixed-sex, secondary, day- or mixed day/boarding, and had at least minimum water sanitation and hygiene (WASH) facilities (an improved water source and functional sex-specific sanitation facilities accessible to female students), and an estimated enrolment as of January 2020 of 50-125 or 40-125 female students in Secondary 1 (first year of secondary school) in Wakiso and Kalungu district, respectively. We selected a random sample of 60 schools confirmed eligible and willing to participate, stratified by government versus private schools and district.

### 2.4 Participant recruitment and informed consent/assent

All female students enrolled in Secondary 2 at the start of the 2022 were eligible to participate in the MENISCUS trial. We sought school-level written consent from headteachers and written informed consent from parents/guardians of students aged <18 years, and from students aged ≥18 years. Students aged <18 years were asked to assent if we had received parental consent.

### 2.5 Data collection

Baseline data were collected from 21 March to 5 July 2022 and endline data were collected from 5 June to 22 August 2023. The surveys included items on sociodemographic characteristics, mental health and multiple dimensions of menstrual health including school absenteeism due to menstruation (“menstrual-related absenteeism”) or due to other reasons, and were self-completed using Open Data Kit software on tablets. The examinations were independently administered and marked by the Uganda National Examination Board (UNEB), with examinations administered on 16-17 March 2022 (baseline) and 24-26 July 2023 (endline). Baseline examinations comprised materials taught in Secondary 1 and included four Mathematics and six Biology questions, of which one question was on puberty and reproduction. Endline examinations additionally included four English Language questions, and comprised materials taught in Secondary 1 and Secondary 2.

### 2.6 Study outcomes

Baseline menstrual-related absenteeism was assessed as self-reported number of school days missed due to menstruation, excluding days where the main reason for missing school was unrelated to menstruation (“Since schools reopened in January this year, how many days of class did you miss during your period?” “What is the main reason for missing school during your period?”). The days of school missed for any reason was obtained from the question (“Since schools reopened in January this year, how many days of class did you miss for any reason?”). We calculated the number of days in the denominator from the official date schools re-opened post COVID-19 (10 January 2022) to the date of interview for each participant, excluding school holidays. Endline menstrual-related absenteeism was assessed as self-reported number of school days missed during menstruation in Terms 1 and 2 of 2023 until the interview date. Term 1 was from 6 February to 5 May 2023, and Term 2 started on 29 May 2023. Baseline and endline examination scores were calculated as the mean of subject-specific z-scores at each time point.

### 2.7 Statistical analyses

Data were exported to Stata version 18 for final cleaning and analysis. We used a hierarchical conceptual framework based on the integrated model of menstrual experience (Hennegan et al 2019) to hypothesize relationships of baseline menstrual-related factors with baseline and endline school absenteeism and educational performance and guide adjustment for confounders. (Figure 1: A conceptual framework show relationship of multiple dimensions of menstrual health with school absenteeism and education performance). In line with the conceptual framework, we defined variables as being in Level 1 (most distal - socio-demographic variables), Level 2 (social support during menstruation), Level 3 (menstrual knowledge, attitudes and adequate menstrual product use), Level 4 (menstrual pain management, menstrual practice needs, experience of menstrual-related teasing), Level 5 (Self-efficacy in addressing menstrual needs scale [SAMNS](20) -menstrual care confidence with menstrual preparedness and pain management sub-scales) and Level 6 (most proximal - trouble concentrating in class). Menstrual-related factors are defined in Table 1.

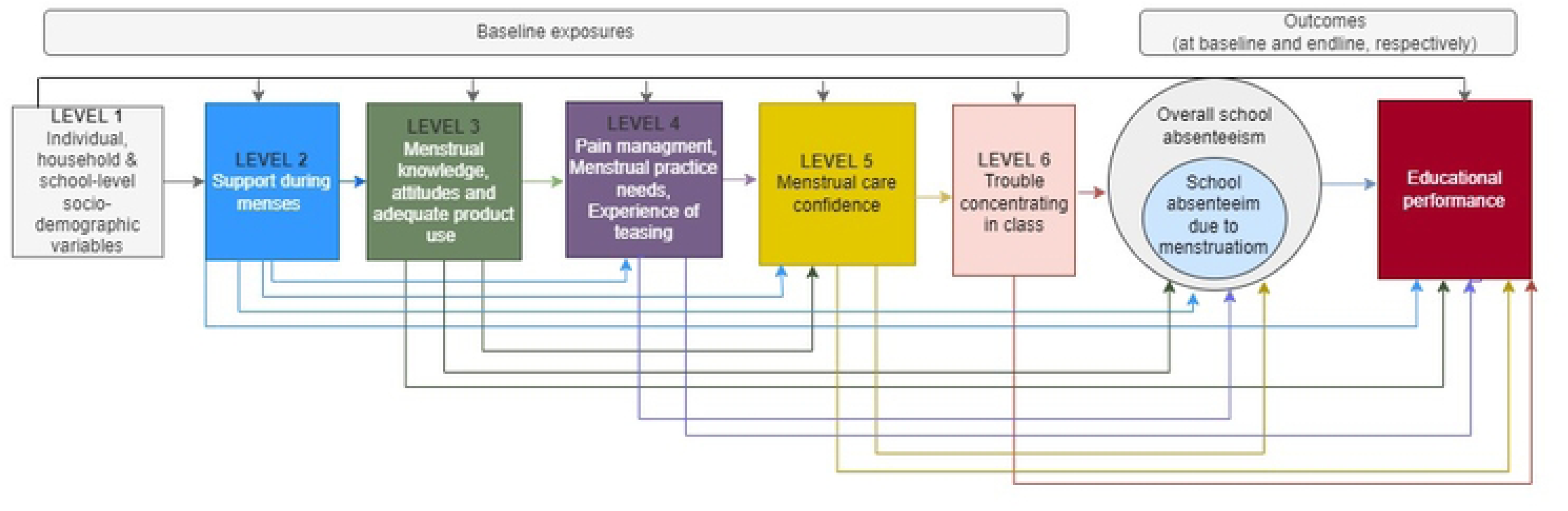

**Table 1:** Definitions of menstrual-related exposure factors used in the conceptual framework.

| Level | Survey measure | Brief description | Calculation | Range & interpretation |
| --- | --- | --- | --- | --- |
| 2 | Social support for menstruation | An individual has someone who they feel okay asking for support for your period if needed (for advice, resources, emotional support) | Binary | Binary (yes/no) |
| 3 | Adequate product use | Used at least one adequate menstrual material and no inadequate materials at LMP | Binary | Binary (yes/no) |
| 3 | Knowledge of puberty & menstruation | Factual knowledge about puberty, menstruation, and the menstrual cycle | Number of 9 knowledge questions answered correctly | 0-9<br>(number of questions answered correctly) |
| 3 | Attitudes towards menstruation | Attitudes about what it is okay for people to do while menstruating and myths about painkillers | Number of 3 attitude questions answered positively | 0-3<br>(number of questions answered positively) |
| 4 | Pain at LMP | Reported having any pain at LMP. This includes reporting any pain, or a symptom of headache, backache, stomach ache or cramp at LMP. | Binary | Binary (yes/no) |
| 4 | Effective pain management strategy | Effective pain strategies defined as stretching, painkillers, eating foods with lots of water, exercising, drinking lots of clean water, holding a warm water bottle on the stomach. Ineffective methods defined as: doing nothing, taking antibiotics, eating spicy foods, drinking soda | Using at least one effective method and no ineffective methods, among girls reporting pain at LMP. | Binary (yes/no) |
| 4 | Menstrual Practice Needs Scale (MPNS-36)(21) | The extent to which individuals' menstrual management practices and environments were perceived to meet their needs during their LMP. | Weighted average of i) core items and school-specific items (75% weight) and ii) relevant material-specific items | Range 0-3. A higher score indicates fewer unmet menstrual needs (i.e. better than a lower score). |

| (25% weight). |  |  |  |  |
| --- | --- | --- | --- | --- |
| 4 | Pain relief | The amount of pain reduced | Binary | All/most or some/none |
| 4 | Experience of teasing | Whether boys or girls tease the participant about their period | Binary | Binary (yes/no) |
| 5 | Self-efficacy in addressing menstrual needs scale (SAMNS-26) (20) | Individuals' confidence in their capabilities to address their menstrual needs. This SAMNS scale has 3 sub-scales 1) The 17-item Menstrual Hygiene Preparation and Maintenance (MHPM) sub-scale includes items regarding self-efficacy in preparing for menstruation (e.g., tracking one's cycle and anticipating days of bleeding), accomplishing various tasks related to maintaining menstrual hygiene (e.g., obtaining, using, cleaning, and disposing, menstrual materials in a variety of contexts), and seeking assistance when needed 2) the 4-item Executing Stigmatized Tasks (EST) sub-scale, which also included items related to obtaining and disposing menstrual materials and seeking assistance—but in situations where accomplishing the tasks involves greater risk of revealing one's menstrual status to a male person and 3) the 5-item Menstrual Pain Management (MPM), measured confidence in mitigating menstrual pain | Mean score | 0-100, representing the mean percentage confidence in addressing needs. Higher % is greater confidence |
| 6 | Trouble participating in class | Whether the participant had trouble participating in class due to period, during their most recent LMP at school | Binary | Binary (yes/no) |

For baseline analysis, we first estimated the mean number of days missed overall per month and the mean number of days reported to be missed due to menstruation. We then used negative binomial regression models to estimate adjusted incidence rate ratios (aIRR) and 95% confidence intervals (CI) for the association of exposures with school absenteeism due to menstruation, allowing for over-dispersion. We accounted for school-level clustering by using random-effects models. Similarly, we used linear regression models to estimate adjusted standardized mean differences (aSMD) and 95%CIs for the association of exposures with examination score, using random-effects to account for school-level clustering. Initially, minimally-adjusted models were fitted for each outcome, adjusting for school-level clustering. We then fitted multivariable models adjusting each variable for other variables at the same or at more distal levels (Figure 1), with the exception of the SAMNS sub-scales, which were not adjusted for each other. We used Wald tests to assess the strength of evidence and refer to a p-value<0.001 as indicating strong evidence of association (22). We interpreted the results in conjunction with effect sizes and confidence intervals to provide comprehensive assessment of results.

To assess temporal associations of exposures with the outcomes, we also analysed associations of baseline exposures with endline outcomes, using analogous methods. We restricted these analyses to participants in the control arm to avoid possible moderating effects of the intervention.

## 3. RESULTS

Of the 4281 eligible participants, 3878 (90.5%) consented/assented and completed the baseline survey. Of these, 3312 (79.1%) participants reported menstruating in the past 6 months and were included in the baseline analyses on school absenteeism. A further 367 (11.1%) participants were not present for the baseline UNEB exam and were excluded from baseline analyses of educational performance. Students who were missing baseline education performance data were more likely to have greater unmet menstrual needs (MPNS score, p=0.01) but there was no evidence of association with other dimensions of menstrual health.

### 3.1 Baseline characteristics of study population

The mean age of participants was 15.6 years (SD=0.9), with most (n=2161; 65.2%) attending a privately-owned school. Around half (n=2070; 55.3%) were day students and had their mother as the primary caregiver (n=1816; 54.8%). A total of 1227 (37.0%) participants lived in households of ≥8 members and 571 (17.2%) reported having one or fewer meals the previous day (Table 2). Overall, 2780 (83.9%) participants reported having social support during their last menstrual period (LMP) and 3038 (91.7%) reported exclusive use of adequate menstrual materials at LMP (Table 3). A total of 1696 (51.2%) participants answered at least 2 of the 3 attitude questions positively, and 538 (16.2%) answered at least 7 of the 9 menstrual knowledge questions correctly. Most participants (2446; 73.9%) reported experiencing menstrual pain at last menstrual period, of whom 1529 (62.5%) reported using an effective pain management strategy. The mean MPNS score was 2.11 (SD=0.52), and participants have a moderate level of menstrual care confidence regarding ability to manage menstrual needs (mean=60.8, SD=19.0). Overall, 251 (7.6%) and 284 (8.6%) participants reported being teased by boys and girls about menstrual periods respectively, and almost half of participants reported trouble participating in class during periods (n=1414, 42.7%) (Table 3).

**Table 2.** Socio-demographic characteristics associated with school absenteeism due to menstruation and examination performance at baseline.

| Variable | Number of participants | Days of school missed due to menstruation per month (n= 3312) | Examination performance (n=2945) |  |  |
| --- | --- | --- | --- | --- | --- |
|  | Frequency (%) | Mean day (SE) | aIRR (95%CI)<sup>1</sup> | Standardized mean exam score (SE) | aSMD (95%CI)<sup>1</sup> |
| **Structural-level characteristics** |  |  |  |  |  |
| **District** |  |  | P=0.38 |  | P=0.23 |
| Wakiso | 2600 (78.5) | 0.29 (0.02) | 1 | 0.03 (0.05) | 0 |
| Kalungu | 712 (21.5) | 0.37 (0.03) | 1.13 (0.86, 1.50) | -0.12 (0.10) | -0.09 (-0.23, 0.05) |
| **School-level characteristics** |  |  |  |  |  |
| **School ownership** |  |  | P=0.80 |  | P=0.99 |
| Private | 2161 (65.2) | 0.27 (0.02) | 1 | 0.07 (0.06) | 0 |
| Government | 1151 (34.8) | 0.38 (0.03) | 0.97 (0.74, 1.27) | -0.16 (0.05) | 0.00 (-0.14, 0.14) |
| **School-mean combined UNEB Math + Biology score** |  |  | P=0.05 |  | P<0.001 |
| High UNEB | 1983 (53.0) | 0.25 (0.02) | 1 | 0.24 (0.05) | 0 |
| Low UNEB | 1758 (47.0) | 0.37 (0.02) | 1.26 (1.00, 1.60) | -0.24 (0.03) | -0.44 (-0.57, -0.32) |
| **Proportion of students boarding at school** |  |  | P=0.95 |  | P=0.84 |
| < 50% | 1820 (55.0) | 0.35 (0.02) | 1 | -0.10 (0.06) | 0 |
| >=50% | 1492 (45.0) | 0.26 (0.02) | 0.99 (0.77, 1.28) | 0.12 (0.05) | 0.01 (-0.12, 0.15) |
| **School size (female baseline participants per school)** |  |  | P=0.57 |  | P=0.24 |
| Greater or equal to 59 | 2258 (68.2) | 0.30 (0.02) | 1 | 0.03 (0.06) | -0.07 (-0.20, 0.05) |
| Fewer than 59 | 1054 (31.8) | 0.34 (0.02) | 0.93 (0.74, 1.18) | -0.04 (0.06) | 0 |
| **Student type** |  |  | P<0.001 |  | P=0.001 |
| Boarding | 1496 (45.2) | 0.24 (0.02) | 1 | 0.11 (0.03) | 0 |
| Day | 1816 (54.8) | 0.37 (0.02) | 1.45 (1.20, 1.77) | -0.07 (0.05) | -0.12 (-0.18, -0.05) |

| Individual-level socio-demographic & economic factors |  |  |  |  |  |
| --- | --- | --- | --- | --- | --- |
| Age group |  |  | P-trend=0.004 |  | P-trend=0.003 |
| <15 | 283 (8.5) | 0.22 (0.03) | 1 | 0.19 (0.07) | 0.00 |
| 15 | 1353 (40.9) | 0.25 (0.02) | 1.15 (0.83, 1.60) | 0.11 (0.05) | -0.02 (-0.13, 0.08) |
| 16 | 1234 (37.3) | 0.36 (0.02) | 1.54 (1.10, 2.15) | -0.08 (0.04) | -0.17 (-0.28, -0.06) |
| 17 | 341 (10.3) | 0.38 (0.05) | 1.66 (1.11, 2.49) | -0.14 (0.06) | -0.23 (-0.36, -0.09) |
| 18+ | 101 (3.0) | 0.48 (0.12) | 1.71 (0.97, 3.03) | -0.35 (0.10) | -0.34 (-0.53, -0.15) |
| Trend |  |  | 1.20 (1.09, 1.33) |  | -0.10 (-0.13, -0.06) |
| Age at first menstruation |  |  | P=0.02 |  | P-trend=0.003 |
| <13 years | 758 (22.9) | 0.34 (0.03) | 1 | 0.09 (0.06) | 0.00 |
| 13 years | 1230 (37.1) | 0.24 (0.02) | 0.74 (0.59, 0.92) | 0.05 (0.05) | -0.01 (-0.08, 0.06) |
| 14 years | 1027 (31.0) | 0.34 (0.03) | 0.92 (0.73, 1.16) | -0.07 (0.05) | -0.08 (-0.16, -0.00) |
| >14 years | 297 (9.0) | 0.40 (0.05) | 1.02 (0.73, 1.42) | -0.20 (0.06) | -0.14 (-0.25, -0.03) |
| Trend |  |  | n/a |  | -0.05 (-0.08, -0.02) |
| Religion |  |  | P=0.002 |  | P=0.19 |
| Catholic | 1052 (31.8) | 0.28 (0.02) | 1 | -0.03 (0.05) | 0 |
| Protestant/SDA | 1285 (38.8) | 0.29 (0.02) | 1.09 (0.88, 1.35) | 0.03 (0.05) | 0.06 (-0.01, 0.13) |
| Muslim | 959 (29.0) | 0.36 (0.03) | 1.45 (1.14, 1.84) | -0.05 (0.05) | -0.01 (-0.09, 0.06) |
| None/Other | 16 (0.5) | 0.87 (0.44) | 3.78 (1.22, 11.74) | 0.03 (0.27) | 0.07 (-0.38, 0.53) |
| Ethnicity |  |  | P=0.54 |  | P=0.83 |
| Muganda | 2280 (68.8) | 0.32 (0.02) | 1 | 0.00 (0.05) | 0 |
| Non Muganda | 1032 (31.2) | 0.30 (0.02) | 0.94 (0.79, 1.14) | 0.02 (0.04) | 0.01 (-0.05, 0.07) |
| Primary caregiver |  |  | P=0.17 |  | P=0.22 |
| Mother | 1938 (58.5) | 0.33 (0.02) | 1 | 0.00 (0.05) | 0 |
| Father | 805 (24.3) | 0.27 (0.02) | 0.85 (0.69, 1.05) | 0.06 (0.05) | 0.04 (-0.02, 0.11) |
| Self and others | 569 (17.2) | 0.29 (0.03) | 0.84 (0.67, 1.06) | -0.10 (0.05) | -0.03 (-0.11, 0.05) |
| Education of primary caregiver |  |  | P=0.15 |  | P=0.001 |
| Secondary or more | 2215 (59.2) | 0.29 (0.02) | 1 | -0.03 (0.05) | 0 |
| Primary or less | 881 (23.6) | 0.38 (0.03) | 1.09 (0.88, 1.34) | 0.04 (0.06) | 0.12 (0.05, 0.19) |
| Don't know | 645 (17.2) | 0.28 (0.03) | 0.83 (0.66, 1.05) | 0.05 (0.05) | 0.09 (0.02, 0.17) |
| **Household size** |  |  | P=0.19 |  | P=0.89 |
| 0-5 | 1001 (30.2) | 0.28 (0.02) | 1 | -0.007 (0.05) | -0.01 (-0.08, 0.05) |
| 6-7 | 1084 (32.7) | 0.31 (0.03) | 1.20 (0.97, 1.49) | -0.001 (0.05) | -0.01 (-0.08, 0.05) |
| >=8 | 1227 (37.0) | 0.34 (0.03) | 1.18 (0.95, 1.45) | 0.00 (0.04) | 0 |
| **Number of meals eaten on the previous day** |  |  | P-trend=0.008 |  | P-trend=0.005 |
| Three or more | 1058 (31.9) | 0.24 (0.02) | 1 | 0.05 (0.05) | 0 |
| Two | 1683 (50.8) | 0.31 (0.02) | 1.20 (0.99, 1.46) | 0.001 (0.05) | -0.03 (-0.09, 0.04) |
| One or fewer | 571 (17.2) | 0.41 (0.04) | 1.53 (1.19, 1.97) | -0.08 (0.05) | -0.13 (-0.21, -0.04) |
| Trend |  |  | 1.24 (1.09, 1.40) |  | -0.06 (-0.10, -0.02) |
| **Socioeconomic status quintile** |  |  | P=0.02 |  | P=0.04 |
| Highest | 656 (19.8) | 0.24 (0.03) | 1 | 0.005 (0.06) | 0 |
| Medium-high | 684 (20.7) | 0.25 (0.03) | 0.91 (0.69, 1.19) | 0.14 (0.04) | 0.12 (0.04, 0.21) |
| Medium | 655 (19.8) | 0.29 (0.03) | 0.99 (0.75, 1.30) | 0.009 (0.06) | 0.04 (-0.05, 0.13) |
| Low-medium | 676 (20.4) | 0.30 (0.03) | 0.92 (0.69, 1.22) | -0.02 (0.05) | 0.03 (-0.06, 0.13) |
| Low | 641 (19.4) | 0.47 (0.04) | 1.35 (1.01, 1.81) | -0.06 (0.05) | 0.00 (-0.09, 0.10) |
| <sup>1</sup> Adjusted for all socio-demographic factors and school level clustering |  |  |  |  |  |

**Table 3.**
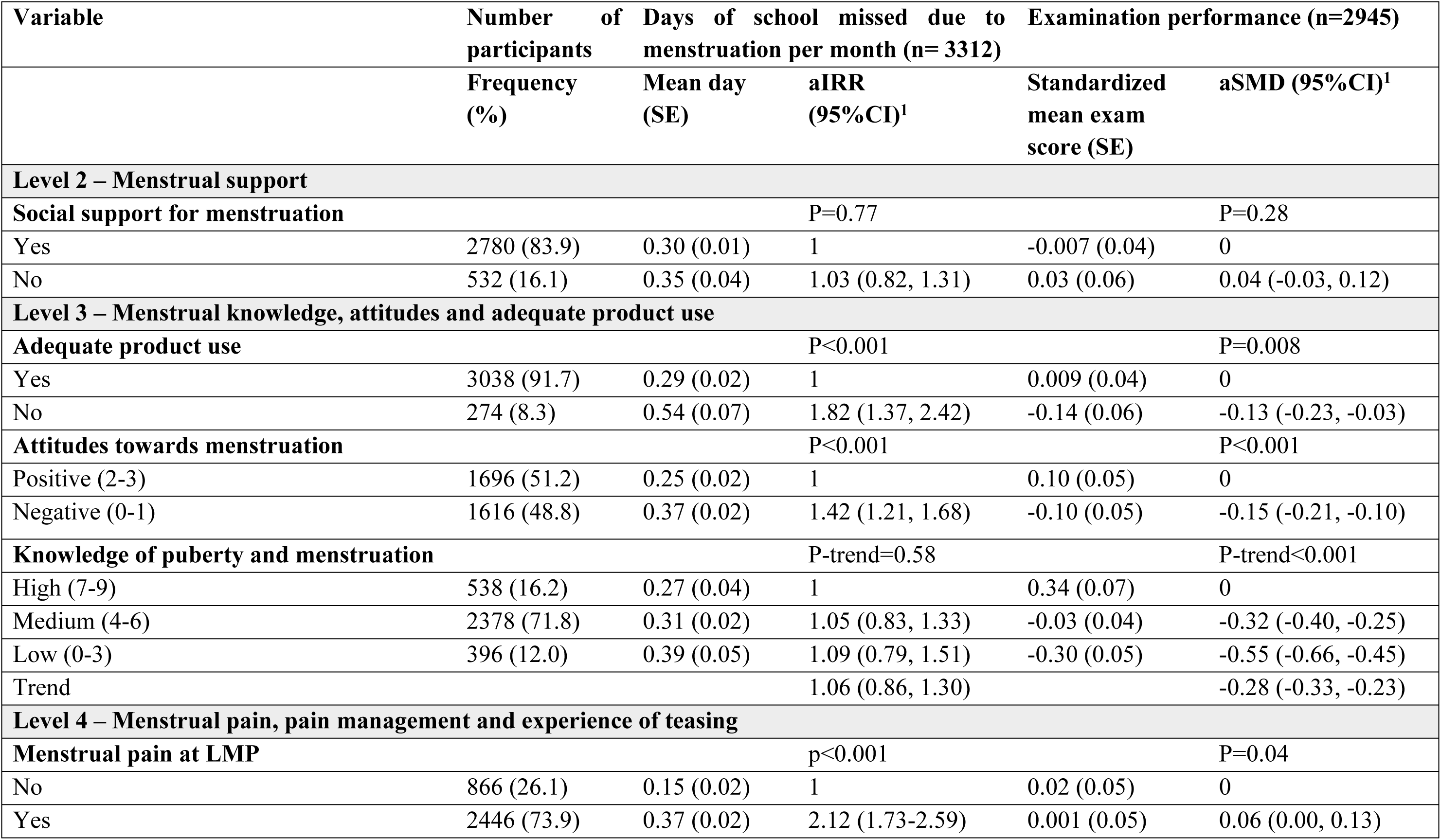
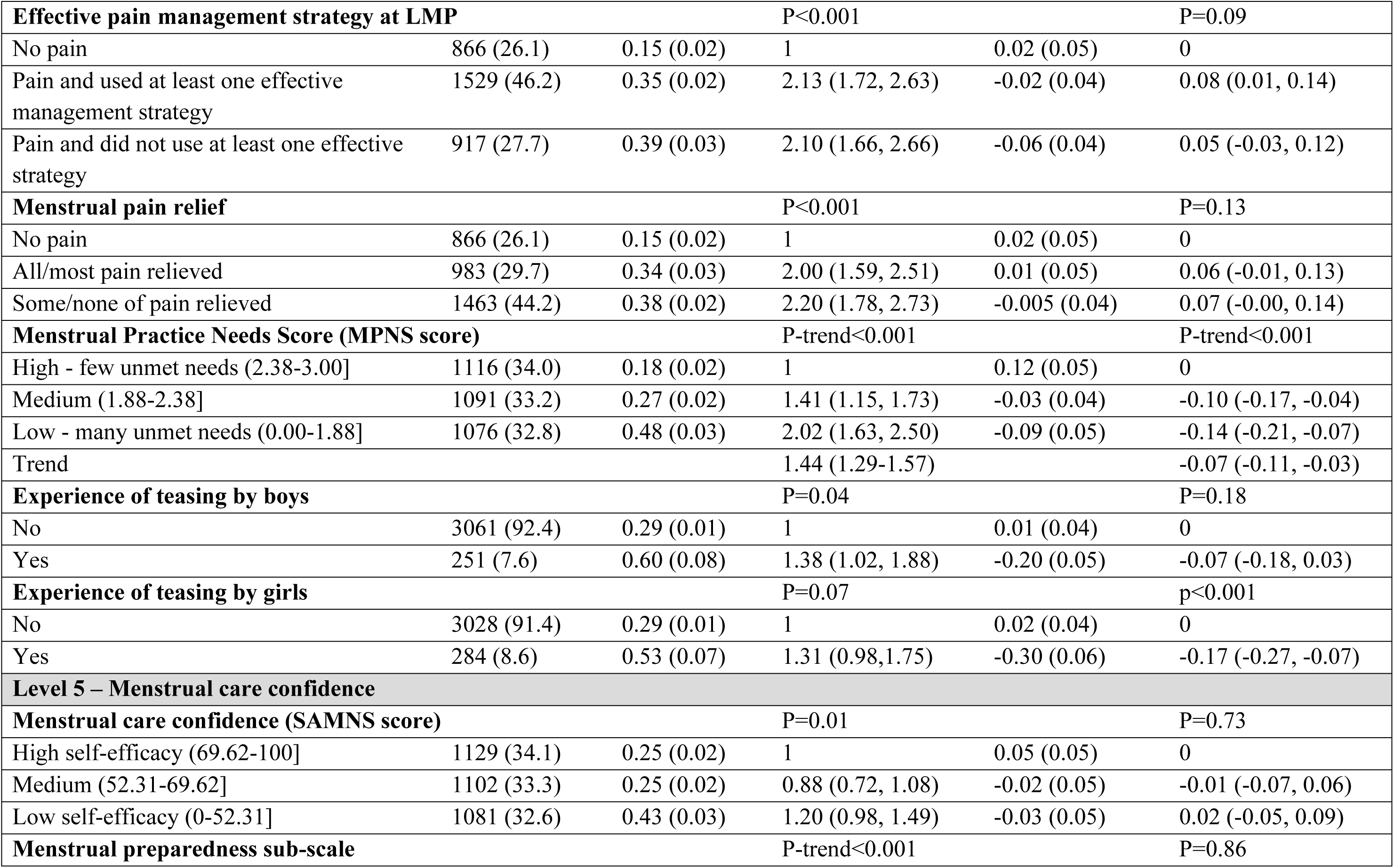
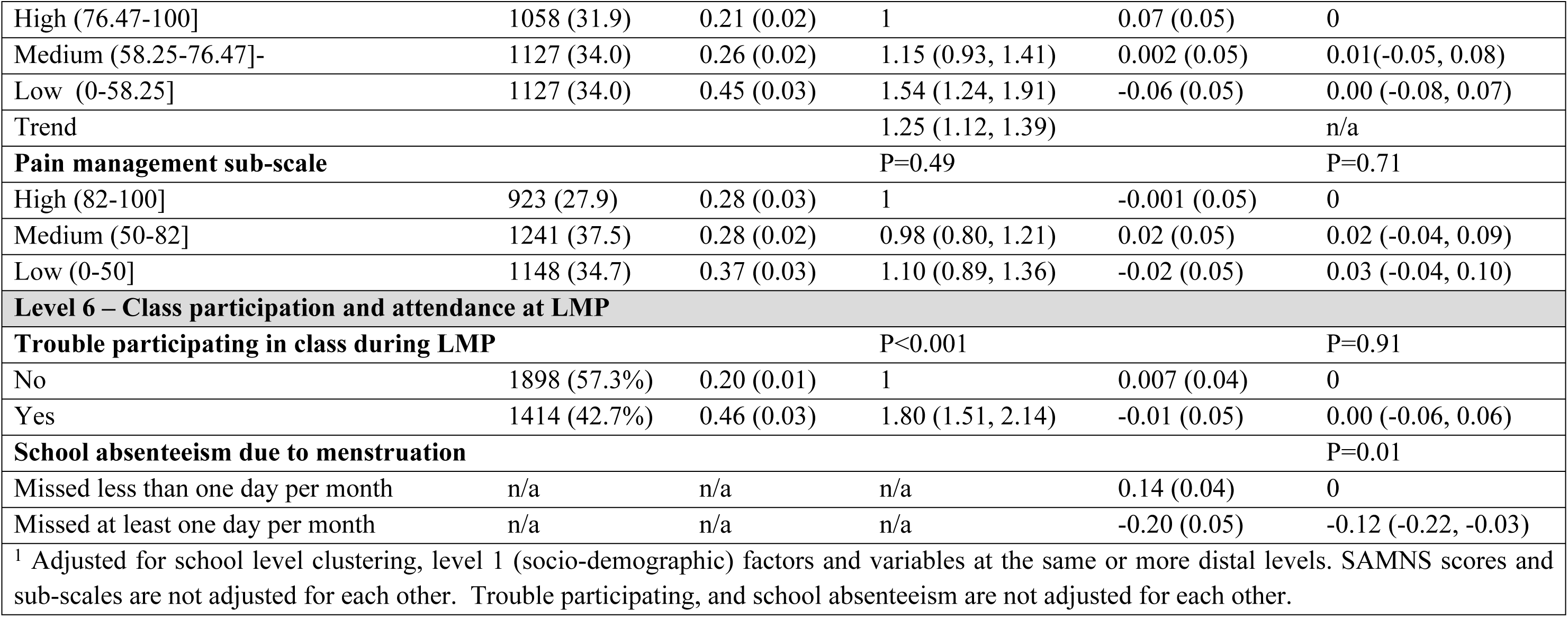
Menstrual characteristics associated with school absenteeism due to menstruation and examination performance at baseline.

| Variable | Number of participants | Days of school missed due to menstruation per month (n= 3312) | Examination performance (n=2945) |  |  |
| --- | --- | --- | --- | --- | --- |
|  | Frequency (%) | Mean day (SE) | aIRR (95%CI)<sup>1</sup> | Standardized mean exam score (SE) | aSMD (95%CI)<sup>1</sup> |
| Level 2 – Menstrual support |  |  |  |  |  |
| Social support for menstruation |  |  | P=0.77 |  | P=0.28 |
| Yes | 2780 (83.9) | 0.30 (0.01) | 1 | -0.007 (0.04) | 0 |
| No | 532 (16.1) | 0.35 (0.04) | 1.03 (0.82, 1.31) | 0.03 (0.06) | 0.04 (-0.03, 0.12) |
| Level 3 – Menstrual knowledge, attitudes and adequate product use |  |  |  |  |  |
| Adequate product use |  |  | P<0.001 |  | P=0.008 |
| Yes | 3038 (91.7) | 0.29 (0.02) | 1 | 0.009 (0.04) | 0 |
| No | 274 (8.3) | 0.54 (0.07) | 1.82 (1.37, 2.42) | -0.14 (0.06) | -0.13 (-0.23, -0.03) |
| Attitudes towards menstruation |  |  | P<0.001 |  | P<0.001 |
| Positive (2-3) | 1696 (51.2) | 0.25 (0.02) | 1 | 0.10 (0.05) | 0 |
| Negative (0-1) | 1616 (48.8) | 0.37 (0.02) | 1.42 (1.21, 1.68) | -0.10 (0.05) | -0.15 (-0.21, -0.10) |
| Knowledge of puberty and menstruation |  |  | P-trend=0.58 |  | P-trend<0.001 |
| High (7-9) | 538 (16.2) | 0.27 (0.04) | 1 | 0.34 (0.07) | 0 |
| Medium (4-6) | 2378 (71.8) | 0.31 (0.02) | 1.05 (0.83, 1.33) | -0.03 (0.04) | -0.32 (-0.40, -0.25) |
| Low (0-3) | 396 (12.0) | 0.39 (0.05) | 1.09 (0.79, 1.51) | -0.30 (0.05) | -0.55 (-0.66, -0.45) |
| Trend |  |  | 1.06 (0.86, 1.30) |  | -0.28 (-0.33, -0.23) |
| Level 4 – Menstrual pain, pain management and experience of teasing |  |  |  |  |  |
| Menstrual pain at LMP |  |  | p<0.001 |  | P=0.04 |
| No | 866 (26.1) | 0.15 (0.02) | 1 | 0.02 (0.05) | 0 |
| Yes | 2446 (73.9) | 0.37 (0.02) | 2.12 (1.73-2.59) | 0.001 (0.05) | 0.06 (0.00, 0.13) |
| **Effective pain management strategy at LMP** |  |  | P<0.001 |  | P=0.09 |
| No pain | 866 (26.1) | 0.15 (0.02) | 1 | 0.02 (0.05) | 0 |
| Pain and used at least one effective management strategy | 1529 (46.2) | 0.35 (0.02) | 2.13 (1.72, 2.63) | -0.02 (0.04) | 0.08 (0.01, 0.14) |
| Pain and did not use at least one effective strategy | 917 (27.7) | 0.39 (0.03) | 2.10 (1.66, 2.66) | -0.06 (0.04) | 0.05 (-0.03, 0.12) |
| **Menstrual pain relief** |  |  | P<0.001 |  | P=0.13 |
| No pain | 866 (26.1) | 0.15 (0.02) | 1 | 0.02 (0.05) | 0 |
| All/most pain relieved | 983 (29.7) | 0.34 (0.03) | 2.00 (1.59, 2.51) | 0.01 (0.05) | 0.06 (-0.01, 0.13) |
| Some/none of pain relieved | 1463 (44.2) | 0.38 (0.02) | 2.20 (1.78, 2.73) | -0.005 (0.04) | 0.07 (-0.00, 0.14) |
| **Menstrual Practice Needs Score (MPNS score)** |  |  | P-trend<0.001 |  | P-trend<0.001 |
| High - few unmet needs (2.38-3.00] | 1116 (34.0) | 0.18 (0.02) | 1 | 0.12 (0.05) | 0 |
| Medium (1.88-2.38] | 1091 (33.2) | 0.27 (0.02) | 1.41 (1.15, 1.73) | -0.03 (0.04) | -0.10 (-0.17, -0.04) |
| Low - many unmet needs (0.00-1.88] | 1076 (32.8) | 0.48 (0.03) | 2.02 (1.63, 2.50) | -0.09 (0.05) | -0.14 (-0.21, -0.07) |
| Trend |  |  | 1.44 (1.29-1.57) |  | -0.07 (-0.11, -0.03) |
| **Experience of teasing by boys** |  |  | P=0.04 |  | P=0.18 |
| No | 3061 (92.4) | 0.29 (0.01) | 1 | 0.01 (0.04) | 0 |
| Yes | 251 (7.6) | 0.60 (0.08) | 1.38 (1.02, 1.88) | -0.20 (0.05) | -0.07 (-0.18, 0.03) |
| **Experience of teasing by girls** |  |  | P=0.07 |  | p<0.001 |
| No | 3028 (91.4) | 0.29 (0.01) | 1 | 0.02 (0.04) | 0 |
| Yes | 284 (8.6) | 0.53 (0.07) | 1.31 (0.98, 1.75) | -0.30 (0.06) | -0.17 (-0.27, -0.07) |
| **Level 5 – Menstrual care confidence** |  |  |  |  |  |
| **Menstrual care confidence (SAMNS score)** |  |  | P=0.01 |  | P=0.73 |
| High self-efficacy (69.62-100] | 1129 (34.1) | 0.25 (0.02) | 1 | 0.05 (0.05) | 0 |
| Medium (52.31-69.62] | 1102 (33.3) | 0.25 (0.02) | 0.88 (0.72, 1.08) | -0.02 (0.05) | -0.01 (-0.07, 0.06) |
| Low self-efficacy (0-52.31] | 1081 (32.6) | 0.43 (0.03) | 1.20 (0.98, 1.49) | -0.03 (0.05) | 0.02 (-0.05, 0.09) |
| **Menstrual preparedness sub-scale** |  |  | P-trend<0.001 |  | P=0.86 |
| High (76.47-100] | 1058 (31.9) | 0.21 (0.02) | 1 | 0.07 (0.05) | 0 |
| Medium (58.25-76.47]- | 1127 (34.0) | 0.26 (0.02) | 1.15 (0.93, 1.41) | 0.002 (0.05) | 0.01(-0.05, 0.08) |
| Low (0-58.25] | 1127 (34.0) | 0.45 (0.03) | 1.54 (1.24, 1.91) | -0.06 (0.05) | 0.00 (-0.08, 0.07) |
| Trend |  |  | 1.25 (1.12, 1.39) |  | n/a |
| **Pain management sub-scale** |  |  | P=0.49 |  | P=0.71 |
| High (82-100] | 923 (27.9) | 0.28 (0.03) | 1 | -0.001 (0.05) | 0 |
| Medium (50-82] | 1241 (37.5) | 0.28 (0.02) | 0.98 (0.80, 1.21) | 0.02 (0.05) | 0.02 (-0.04, 0.09) |
| Low (0-50] | 1148 (34.7) | 0.37 (0.03) | 1.10 (0.89, 1.36) | -0.02 (0.05) | 0.03 (-0.04, 0.10) |
| **Level 6 – Class participation and attendance at LMP** |  |  |  |  |  |
| **Trouble participating in class during LMP** |  |  | P<0.001 |  | P=0.91 |
| No | 1898 (57.3%) | 0.20 (0.01) | 1 | 0.007 (0.04) | 0 |
| Yes | 1414 (42.7%) | 0.46 (0.03) | 1.80 (1.51, 2.14) | -0.01 (0.05) | 0.00 (-0.06, 0.06) |
| **School absenteeism due to menstruation** |  |  |  |  | P=0.01 |
| Missed less than one day per month | n/a | n/a | n/a | 0.14 (0.04) | 0 |
| Missed at least one day per month | n/a | n/a | n/a | -0.20 (0.05) | -0.12 (-0.22, -0.03) |
| <sup>1</sup> Adjusted for school level clustering, level 1 (socio-demographic) factors and variables at the same or more distal levels. SAMNS scores and sub-scales are not adjusted for each other. Trouble participating, and school absenteeism are not adjusted for each other. |  |  |  |  |  |

Participants reported missing a mean of 1.25 school days per month overall (95%CI 1.17, 1.33), and 0.30 days due to menstruation (95%CI 0.27, 0.34). A total of 991 (30%) participants reporting missing school due to menstruation since schools reopened, with 323 (9.8%) reporting missing at least one day per month due to menstruation.

### 3.2 Socio-demographic factors associated with menstrual-related absenteeism and examination performance at baseline

Menstrual-related absenteeism and poorer examination performance were both associated with being in a school with a poorer examination performance (aIRR=1.26, 95%CI 1.00, 1.60; aMD=-0.44, 95%CI -0.57, -0.32), being a day student (aIRR=1.45, 95%CI 1.20, 1.77; aMD=-0.12, 95%CI -0.18, -0.05), older age (aIRR-trend=1.20, 95%CI 1.09, 1.33; aMD-trend=-0.10, 95%CI -0.13, -0.06) and having fewer meals the previous day (aIRR-trend=1.24, 95%CI 1.09, 1.40; aMD-trend=-0.06, 95%CI -0.10, -0.02) (Table 2). In addition, poorer examination performance was associated with older age at first menses (aMD-trend=-0.05, 95%CI -0.08, -0.02) and higher education of the primary caregiver (aMD=-0.12, -0.19, -0.05) (Table 2).

### 3.3 Menstrual factors associated with school absenteeism due to menstruation and poorer examination performance at baseline

Among the 991 participants reporting missing school due to menstruation, 1023 days were missed per month (mean per participant =1.03 per month (SE=0.04) (Table 4). The most commonly reported main reason was illness (back/stomach pain or cramps: n=588; 57.2% of days missed; feeling generally unwell: n=170; 17.2% of days missed). Relatively few participants reported other reasons (stigma-related, the physical environment or socio-economic) as the main reason for missing school during menstruation (Table 4).

**Table 4:** Main reason for missing school due to menstruation, reported at baseline.

| **Main reason for missing school due to menstruation** | **Number of participants n (%)** | **Mean (SD) days per participant missed due to menstruation per month among those reporting this reason** | **Total days missed due to menstruation per month with this as the main reason** | **Proportion of days missed due to menstruation per month with this as the mains reason** |
| --- | --- | --- | --- | --- |
| **Total** | **991** | **1.03 (1.16)** | **1023** | **100%** |
| **Illness** |  |  |  |  |
| Back/stomach pain or cramps | 588 (59.3) | 1.00 (1.06) | 585 | 57.2% |
| Feeling generally unwell | 170 (17.2) | 0.98 (1.20) | 165 | 16.2% |
| **Stigma** |  |  |  |  |
| Scared of leaking blood on clothes | 59 (6.0) | 0.92 (0.95) | 54 | 5.3% |
| Not wanting to change pad or absorbents at school | 2 (0.2) | 2.49 (3.01) | 5 | 0.5% |
| Household members telling me not to go to school during period | 12 (1.2) | 1.86 (2.38) | 22 | 2.2% |
| Worried that others may know I have my periods | 18 (1.8) | 0.79 (0.53) | 14 | 1.4% |
| Scared of having to ask for help because of periods | 6 (0.6) | 0.94 (0.72) | 6 | 0.5% |
| **Physical environment** |  |  |  |  |
| School toilets or latrines not clean | 34 (3.4) | 1.20 (1.28) | 41 | 4.0% |
| Lack of access to toilet paper at school | 4 (0.4) | 0.56 (0.24) | 2 | 0.2% |
| Lack of access to water and/or soap at school | 17 (1.7) | 1.34 (2.27) | 23 | 2.2% |
| Not enough privacy in school toilet/latrine | 34 (1.4) | 1.20 (1.28) | 41 | 4.0% |
| **Socio-economic** |  |  |  |  |
| Unable to afford pads or other absorbents | 35 | 1.50 (1.45) | 52 | 5.1% |
| Lack of knickers | 31 | 1.26 (1.35) | 39 | 4.8% |

After adjusting for factors at the same or more distal levels, there was strong evidence that menstrual-related absenteeism and poorer examination performance were both associated with not exclusively using adequate menstrual materials (aIRR=1.82, 95%CI 1.37, 2.42; aMD=-0.13, 95%CI -0.23, -0.03), negative menstrual attitudes (aIRR=1.42, 95%CI 1.21, 1.68, aMD=-0.15, 95%CI -0.21, -0.10), menstrual pain at LMP (aIRR=2.12, 95%CI 1.73, 2.59; aMD=0.06, 95%CI 0.00, 0.13), more unmet menstrual practice needs (p-trend<0.001) and reported experience of menstrual-related teasing at LMP by girls (aIRR=1.31, 95%CI 0.98, 1.75; aMD=-0.17, 95%CI -0.27, -0.07) (Table 3). In absolute terms, the difference was relatively small, for example, the mean days missed per month among participants with no menstrual pain at LMP was 0.15 (SE 0.02), compared with 0.37 (SE 0.02) among participants with pain at LMP.

After adjustment, menstrual-related absenteeism was associated with lack of perceived pain relief at LMP (aIRR=2.20, 95%CI 1.78, 2.73 for minimal/no pain relieved vs no pain), menstrual teasing by boys (aIRR=1.38, 95%CI 1.02, 1.88), poor menstrual care confidence (aIRR=1.20, 95%CI 0.98, 1.49 for lowest vs highest quartile, p-trend=0.01), especially due to lack of menstrual preparedness (aIRR=1.54, 95%CI 1.24, 1.91, p-trend<0.001) and with trouble participating in class during LMP (aIRR=1.80, 95%CI 1.51, 2.14). Similar factors were associated with missing school overall (i.e. for any reason) (Supplementary Table 1).

### 3.4 Menstrual factors at baseline associated with menstrual-related absenteeism and poorer examination performance at endline

Among the 1652 participants in control arm schools at baseline, 1253 (75.8%) were in control schools at endline and completed the endline survey. Of these, 1192 (95.1%) reported having menstruated in the past 6 months and were included in endline analysis, and 1099 (92.2%) had examination data performance at endline.

Among these participants, menstrual-related absenteeism and poorer examination performance at endline were both associated with using inadequate menstrual products at baseline (aIRR=1.76, 95%CI 0.95, 3.24; aMD=-0.19, 95%CI -0.34, -0.03), negative menstrual attitudes (aIRR=1.76, 95%CI 1.26, 2.46; aMD=-0.16, 95%CI -0.24, -0.08), more unmet menstrual practice needs (aIRR=1.75, 95%CI 1.12-2.71; aSMD= -0.10, 95%CI -0.21 to 0.00) and experience of teasing about menstruation by girls (aIRR=1.89, 95%CI 1.06, 2.73; aMD=-0.33, 95%CI -0.48, -0.18) (Table 5).

**Table 5.**
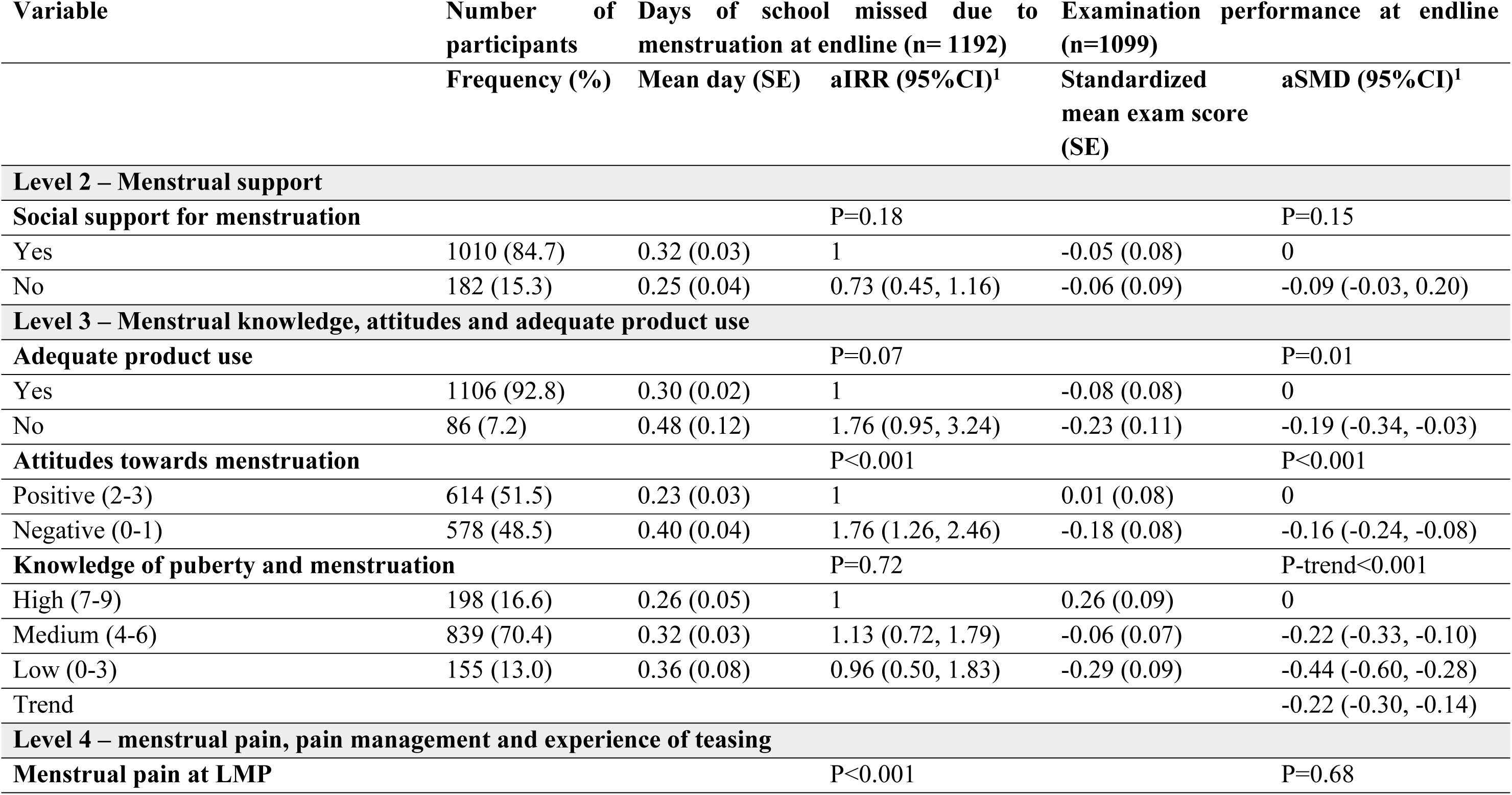
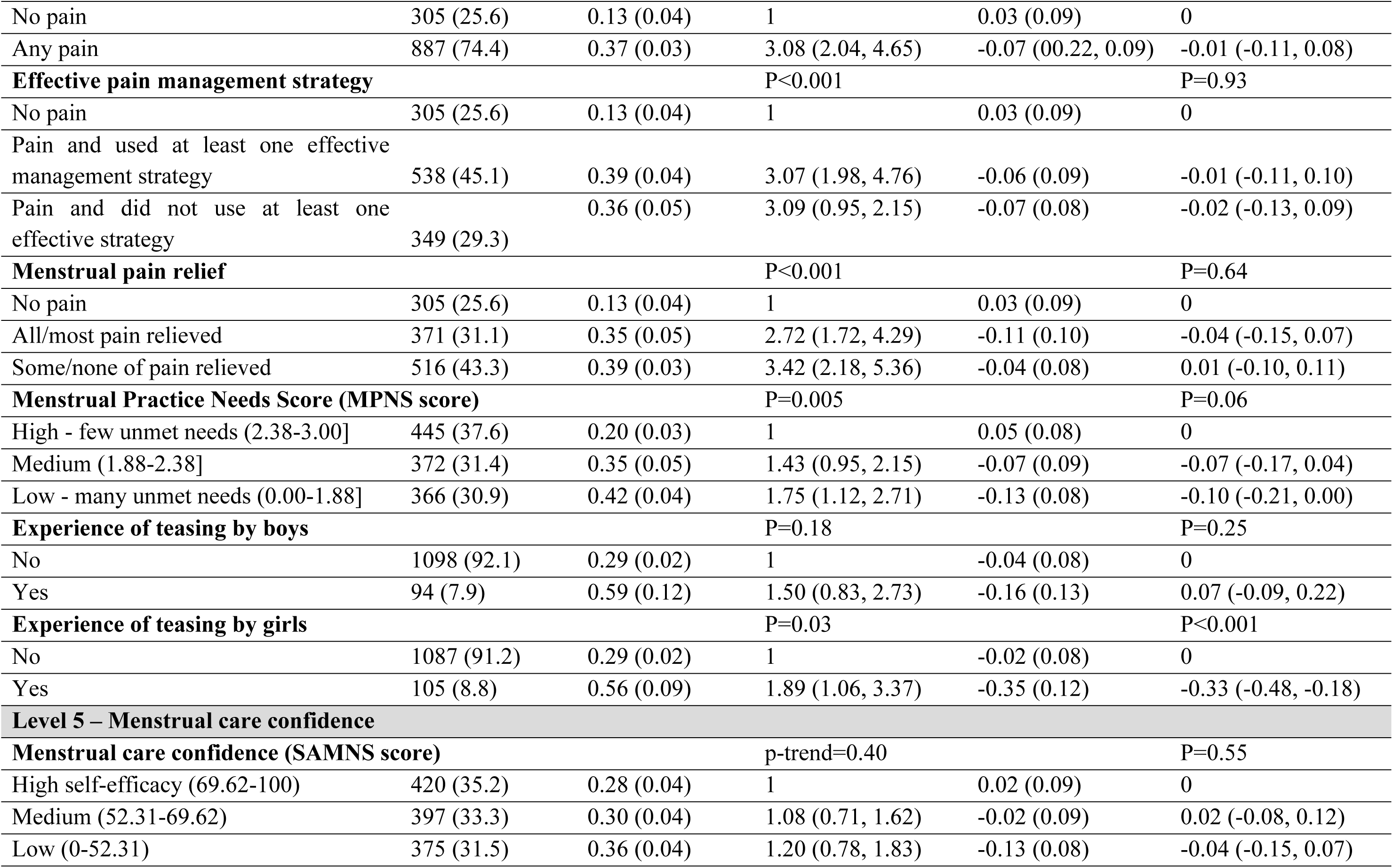
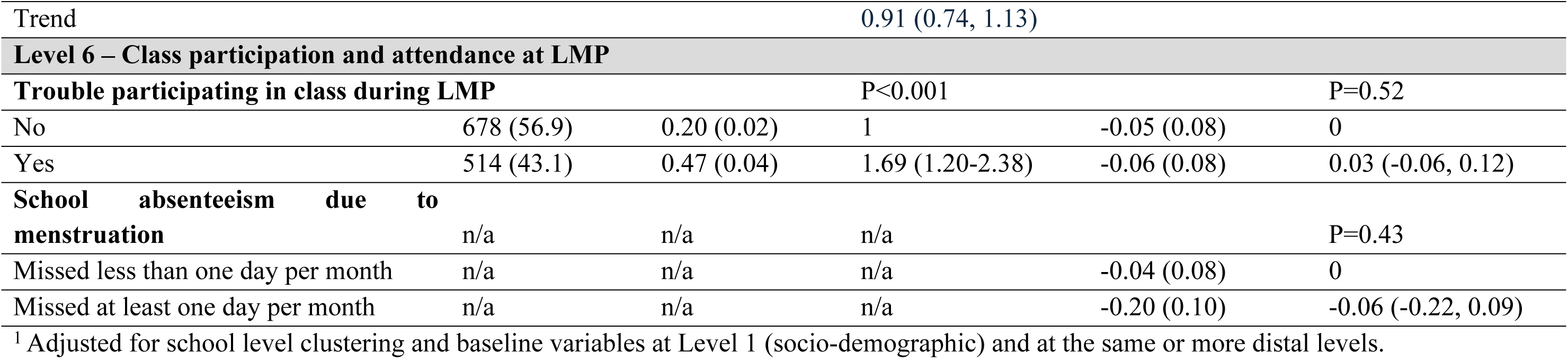
Baseline menstrual characteristics associated with endline school absenteeism due to menstruation and examination performance at baseline.

In addition, menstrual-related absenteeism was associated with baseline menstrual pain (aIRR=3.08, 95%CI 2.04, 4.65), including if there was pain management but no pain relief (aIRR=3.42, 95%CI 2.18, 5.36 vs no pain), and trouble participating in class during LMP (aIRR=1.69, 95%CI 1.20, 2.38). Endline examination performance was additionally associated with poorer baseline menstrual knowledge (aMD=-0.44, 95%CI -0.60, -0.28) (Table 5).

Similar factors were associated with school absenteeism for any reason (including menstruation) (Supplementary Table 1). For example, the mean number of days missed due to menstrual pain was 1.36 days per month, vs 0.88 among those without pain at LMP (aIRR=1.41, 95%CI 1.26-1.57).

## 4. DISCUSSION

### 4.1 Multiple dimensions of menstrual health are associated with school absenteeism and poor examination performance

In this setting, there is strong evidence that multiple dimensions of menstrual health were associated with both increased menstrual-related absenteeism and poorer examination performance. These included use of inadequate products, negative attitudes towards menstruation, having many unmet menstrual needs, menstrual pain and experience of teasing. These add to the limited evidence quantifying both the amount of menstrual-related absenteeism, and the specific menstrual-related factors associated with absenteeism and examination performance.

### 4.2 The impact of menstrual health on number of days of school missed due to menstruation

Our findings that 10% of participants reported missing at least one school day per month due to menstruation supports the widespread claim that one in 10 girls miss school due to menstruation (3). However, on average, participants reported missing less than half a day of school per month due to menstruation (0.30 days). The prevalence of menstrual-related absenteeism is at the lower end of the range seen in the systematic review of studies in Sub-Saharan Africa (6-65% missing school during menstruation) (5), and slightly lower than the recent multi-country study (17.7%, 95%CI 15.1-20.3%) among 15-19 year olds (6). Reported school absence may be lower in our study because i) students had missed almost two years of school due to COVID-19 closures and may have felt pressure from teachers and parents to make up the time; ii) reported days of school missed due to menstruation may be under-reported by boarding students who interpreted being on school premises as attending school, and iii) the relatively long period of reporting in our study may have led to underestimation due to recall bias. The variation in estimates between studies is also likely due to challenges measuring school attendance accurately (8). Measuring school attendance using self-report often underestimates absenteeism compared to using school registers (23), which may be due to social desirability bias (8, 24), stigma and taboos around menstruation, and recall bias. This is supported by qualitative findings from a Kenyan study, in which girls rarely reported school absenteeism themselves, but reported that ‘other’ girls did (25). However in some settings, including Uganda, school registers are incomplete and/or inaccurate, due to teachers not completing the registers, and social desirability bias if school funding is linked with student numbers (23). Future studies should consider a combination of these methods, daily diaries and spot checks to triangulate data on school attendance (23).

In our study, students reporting menstrual pain had more than double the rate of menstrual-related absenteeism than those with no pain, and school absenteeism was highest among those with inadequate pain relief. Further, we found that menstrual pain or feeling generally unwell were responsible for about three-quarters of days of school absenteeism due to menstruation. To our knowledge, this is the first study to quantify days of school absenteeism attributable to specific menstrual-related factors.

At baseline, there was evidence that participants with low menstrual self-efficacy were more likely to miss school due menstruation compared to those with high menstrual self-efficacy. The association was strongest with the sub-scale of lack of menstrual preparedness (e.g., tracking one’s cycle, obtaining, using, cleaning, and disposing of menstrual products, and seeking assistance when needed). This fits with findings from qualitative studies that students with low menstrual confidence might feel uncomfortable to be around students or teachers during menstruation and hence choose to miss school during menstruation (9, 26). This association may also be due to girls fearing leakage of blood whilst in class and hence missing school (2).

### 4.3 The impact of menstrual health on educational performance

Qualitative studies have described how participants perceive that menstruation affects their academic performance (2) but there are few rigorous quantitative studies on this. Our findings support the hypothesis that menstruation affects academic performance through multiple pathways, through menstrual pain or use of inadequate materials, but also due to social stigma and lack of menstrual knowledge. Using a standardised assessment set by the UNEB, we found that poorer examination performance was associated with multiple dimensions of menstrual health. There was strong evidence of associations between poorer examination scores and use of inadequate materials, poorer menstrual attitudes, poorer menstrual knowledge, having more unmet menstrual practice needs and experience of being teased in relation to periods.

Our results support qualitative findings from the systematic review (2) and a qualitative study of 120 girls aged 14-16 years in in Kenya where girls reported that their attention in class was disturbed by worries of leakage of menstrual blood and staining of clothing (27). A further study found that students reported failing to stand up and answer questions, write on the board and miss exams during menstruation (12). Missing at least one day of school due to menstruation at LMP at baseline was associated with poorer baseline examination scores, but not with endline examination scores. This difference may be due to chance or due to the longer temporal gap between missing school at baseline and examination performance at endline. Persistent school absenteeism increases the risk of school dropout, lowers girls’ self-esteem, and compromises future productivity (28). A strength of our study was the use of an independent examination set by UNEB. We identified one previous study in Uganda which evaluated academic performance similarly, using assessments in English language, Mathematics, Integrated Science, and Social Studies set by UNEB (29) . In this small randomised controlled trial of 60 participants in two primary schools in Uganda, a menstrual health intervention (which included telling menstrual stories and playing games related to menstrual management) was associated with improved academic scores at 6 weeks after the intervention (29)

### 4.4 Strengths and limitations of the study

Strengths of our study include the measurement of multiple dimensions of menstrual health; use of both cross-sectional and longitudinal analyses (to minimise reverse causality); measurement of reported number of schooldays missed due to menstruation over a defined period; use of a self-completed questionnaire (to minimise social desirability bias); use of validated tools for menstrual care confidence and unmet practice needs; and assessment of educational performance using an examination set independently by UNEB, tailored to material that had been taught across all 60 schools.

Limitations include possible recall bias from retrospective, self-reported outcome of school attendance, residual confounding, and challenges related to the long duration (one year) between baseline menstrual experiences and endline outcomes for the longitudinal analyses. The educational performance outcome is objective but limited, especially at baseline, by the relatively sparse amount of material that had been taught between schools re-opening after COVID-9 in January 2022, and the assessment in March 2022. This resulted in a lack of discrimination between potentially low- and high-achieving students.

### 5.5 Conclusion and Recommendation

Multiple dimensions of menstrual health are strongly associated with school absenteeism and subsequent poor education performance among female students in Ugandan secondary schools, but the absolute effect size is small as menstrual-related absenteeism is infrequent. Our previous finding that there was no evidence of an effect of a multi-component menstrual health intervention on school absenteeism and educational performance in this population, despite modest improvements in multiple dimensions of menstrual health (15), suggests that larger improvements in menstrual health are needed to impact on educational outcomes. However, improving menstrual health remains a critical priority globally. Addressing this issue is essential not only for safeguarding physical and psychological well-being but also advancing gender equality and human rights issue.

## Data Availability

The data are held in a public repository here: LSHTM Data Compass Weiss, H, Nelson, K, Mugenyi, L, Batuusa, L, Thomas, K and Baleke, C (2024). MENISCUS Trial. [Project]. London School of Hygiene & Tropical Medicine, London, United Kingdom. https://doi.org/10.17037/DATA.00003822.

## Acknowledgements

We would like to thank the staff, students, and communities of participating schools for their engagement with the study; the stakeholders from the Uganda Ministry of Education and Sports; and District Education Officers in Wakiso and Kalungu districts.

## Abbreviations

MH: Menstrual health
LMP: Last menstrual period
CI: Confidence interval
LRT: Likelihood ratio test
SMD: Standardized mean difference
IRR: Incidence rate
MENISCUS: Menstrual health interventions, schooling, and mental health problems among Ugandan students
MHM: Menstrual hygiene management
SES: Socio-economic status
WASH: Water, sanitation and hygiene
SAMNS: Self-efficacy in Addressing Menstrual Needs Scale
MHPM: Menstrual Hygiene Preparation and Maintenance sub-scale
MPM: Menstrual Pain Management sub-scale
EST: Executing Stigmatized Tasks sub-scale

## Declarations Ethical approval

Ethical approval for MENISCUS protocol, the informed consent forms and draft CRFs were obtained from the Uganda Virus Research Institute Research & Ethics Committee (UVRI-REC) (reference GC/127/819) and the Uganda National Council of Science and Technology (UNCST) (reference HS1525ES) and the LSHTM Research Ethics Committee (reference 22952-2).

## Consent for publication

Participants consented to the research findings being published in international science journals and electronic websites, on condition that that findings cannot be traced to individuals.

## Availability of data and materials

The datasets used and/or analysed during the current study are available from the corresponding author on reasonable request.

